# Comparison of Four Molecular *In Vitro* Diagnostic Assays for the Detection of SARS-CoV-2 in Nasopharyngeal Specimens

**DOI:** 10.1101/2020.04.17.20069864

**Authors:** Wei Zhen, Ryhana Manji, Elizabeth Smith, Gregory J. Berry

**Affiliations:** Infectious Disease Diagnostics, Northwell Health Laboratories, Lake Success, NY; Department of Pathology and Laboratory Medicine, The Donald and Barbara Zucker School of Medicine at Hofstra/Northwell

**Keywords:** SARS-CoV-2, COVID-19, EUA, molecular diagnostics, real-time RT-PCR, nasopharyngeal

## Abstract

The novel human coronavirus SARS-CoV-2 was first discovered in the city of Wuhan, Hubei province, China, causing an outbreak of pneumonia in January 2020. As of April 10, 2020, the virus has rapidly disseminated to over 200 countries and territories, causing more than 1.6 million confirmed cases of COVID-19 and 97,000 deaths worldwide. The clinical presentation of COVID-19 is fairly non-specific, and symptoms overlap with other seasonal respiratory infections concurrently circulating in the population. Further, it is estimated that up to 80% of infected individuals experience mild symptoms or are asymptomatic, confounding efforts to reliably diagnose COVID-19 empirically. To support infection control measures, there is an urgent need for rapid and accurate molecular diagnostics to identify COVID-19 positive patients. In the present study, we have evaluated the analytical sensitivity and clinical performance of four SARS-CoV-2 molecular diagnostic assays granted Emergency Use Authorization by the FDA using nasopharyngeal swabs from symptomatic patients. This information is crucial for both laboratories and clinical teams, as decisions on which testing platform to implement are made.

## Introduction

A novel human virus, severe acute respiratory syndrome coronavirus 2 (SARS-CoV-2), was identified as the causative agent for an outbreak of viral pneumonia that began in Wuhan, China at the end of 2019 (1). On March 11, the SARS-CoV-2 epidemic was escalated to the level of a global pandemic by the World Health Organization (WHO) (1). The WHO has named the illness caused by SARS-CoV-2 as coronavirus disease-2019 (COVID-19). COVID-19 has since continued to spread across the globe, and as of April 10, 2020, over 1.6 million cases have been confirmed in more than 200 countries and territories, causing over ∼97,000 deaths. More than ∼460,000 confirmed COVID-19 cases and ∼16,000 deaths have been reported in the United States according to the Centers for Disease Control and Prevention (CDC) and database from the Center for System Science and Engineering (CSSE) at Johns Hopkins University (2, 3).

SARS-CoV-2 (COVID-19) is the seventh coronavirus known to infect humans; SARS-CoV, MERS-CoV and SARS-CoV-2 can cause severe disease, whereas seasonal coronavirus HKU1, NL63, OC43 and 229E are associated with mild symptoms (4). Coronaviruses are a diverse family of large RNA viruses that are known to be involved in zoonotic transmission between a wide variety of animals and humans. Coronaviruses generally target epithelial cells in the respiratory and gastrointestinal tracts, and viral shedding can occur from these sites. Infection caused by coronaviruses can therefore typically be transmitted through several different routes, including aerosol and fecal-to-oral, with fomites often playing an important role in the infection cycle (5). Notably, coronaviruses possess a distinctive morphological feature, a ring of spike proteins on the outer surface of the virus, giving the appearance of a halo or corona. In addition to inspiring the name of the coronavirus genus, the spike proteins are also essential for infection of host cells. The SARS-CoV-2 spike protein recognizes and binds to the human cellular receptor angiotensin-converting enzyme 2 (ACE2), then subsequently mediates fusion of the viral and host cell membranes, allowing the virus to gain entry (6, 7). The ACE2 receptor is found on epithelial cells of the lungs and small intestines (8).

Infection with SARS-CoV-2 can cause mild to severe respiratory illness and symptoms include fever, cough and shortness of breath. However, some populations experience severe, rapidly progressive and fulminant disease. This population includes: older adults and people who have serious underlying medical conditions (e.g. heart disease, diabetes, lung disease and immunosuppression) (9). Unfortunately, many elements, some intrinsic to the virus and others seasonal, have lessened the effectiveness of traditional infection control measures. The combination of high rates of human-to-human transmission (R_0_ = 2.0-2.5), stability of the virus in aerosols and on surfaces, the fairly non-specific clinical presentation of COVID-19, along with co-incidence with the active season of other respiratory viruses (e.g., influenza, respiratory syncytial virus (RSV)) in many parts of the world, together present a major challenge to stop the pandemic from spiraling into a more severe global health emergency (9, 10).

During the early stages of the epidemic, both national and international agencies rushed to initiate the process of mass production of test reagents and issued an Emergency Use Authorization (EUA) for the US CDC COVID-19 real-time RT-PCR assay (11). Despite the collective effort, laboratories are still facing reagent supply shortages, lack of instrument access, an inability to perform high-complexity testing as well as facing increased staffing needs, leaving a gap in the ability of healthcare providers to rapidly diagnose and manage patients. The need to implement a sensitive, accessible, and rapid diagnostic test for the detection of COVID-19 is warranted. In this study, we evaluated the analytical and clinical performance of four SARS-CoV-2 molecular diagnostic assays granted EUA by the FDA, including the modified CDC, DiaSorin Molecular, GenMark, and Hologic assays. These assays are authorized for the qualitative detection of SARS-CoV-2 in clinical specimens obtained from symptomatic patients and were evaluated using nasopharyngeal swab specimens.

## Materials and Methods

### Specimen collection and storage

Sterile nylon, Dacron or rayon swabs with flexible plastic shafts were used to collect nasopharyngeal specimens from each patient. After collection, the swabs were placed into 3 ml of sterile Universal Transport Medium (UTM; various manufacturers). Specimens were tested as soon as possible after collection, or if testing was delayed, they were stored for up to 72 h at 2-8°C after collection. Following routine testing, samples were stored frozen (≤-80°C) until comparator testing could be completed.

### Study design

A total of 104 retrospective and prospective nasopharyngeal specimens originally submitted for routine COVID-19 testing at Northwell Health Laboratories on the GenMark ePlex SARS-CoV-2 panel were selected for this study. Of the 104 specimens analyzed, 51 were positive and 53 were negative samples. Retrospective frozen samples were thawed, pipetted into separate aliquots, and were tested by the modified CDC assay, DiaSorin Molecular Simplexa COVID-19 Direct assay, and Hologic Panther Fusion^®^ SARS-CoV-2 assay in parallel. Manufacturer’s specifications are summarized in **Table 1**. The study population included patients of all ages and both genders presenting with signs and/or symptoms of COVID-19 infection, and low viral loads determined by high cycle threshold (Ct) results generated during routine testing.

**Table 1.** Overview of four molecular *in vitro* diagnostic EUA assays used in this study.

|  | 2019-Novel<br>Coronavirus (2019-nCoV) | Simplexa™ COVID-19<br>Direct | ePlex SARS-CoV-2<br>Test | SARS-CoV-2 Assay |
| --- | --- | --- | --- | --- |
| Manufacturer | Modified CDC | Diasorin Molecular | GenMark | Hologic |
| Sample type | NPS, OPS, Sputum | NPS, NS, BAL | NPS | NPS, OPS |
| Sample volume<br>required (μl) | 110 | 50 | 200 | 500 |
| Extraction required | Yes (manual) | No | Yes (automated) | Yes (automated) |
| Detection<br>platform/System | ABI 7500 Fast Dx | LIAISON® MDX | ePlex® | Panther Fusion® |
| Target region of SARS-CoV-2 | N (N1 & N2) | S and ORF1ab | N | ORF1ab |
| Analytical sensitivity<br>per claim | 500 copies /mL | 500 copies /mL | 100,000 copies /mL | 1x10<sup>-2</sup>TCID<sub>50</sub>/mL |

### The New York SARS-CoV-2 Real-time Reverse Transcriptase (RT)-PCR Diagnostic EUA Panel (Modified CDC assay)

This assay is a modified version of the CDC 2019-Novel Coronavirus (2019-nCoV) Real-Time RT-PCR Diagnostic EUA Panel validated by the Wadsworth Center using the same primer and probe sets as the CDC assay for nucleocapsid (N) gene N1 and N2 targets and human RNase P gene (RP), but excluding the N3 primer and probe set. A volume of 110 µL patient specimen was extracted by the NucliSENS easyMAG platform (BioMérieux, Durham, NC) according to manufacturer’s instructions, with a nucleic acid elution volume of 110 µL. For each specimen, three Master Mix sets including N1, N2, and RNase P were prepared, and 15 µL of each master mix was dispensed into appropriate wells, followed by 5 µL of extracted sample. Each run also included a No Template Control (NTC), Negative Extraction Control and a SARS-CoV-2 Positive Control. Amplification was performed on the Applied Biosystems^®^ 7500 Fast Dx Real-Time PCR System. The results interpretation algorithm for reporting a positive specimen requires both N1 and N2 targets to be detected.

### DiaSorin Molecular Simplexa COVID-19 Direct EUA

Testing with the DiaSorin Molecular Simplexa COVID-19 Direct EUA assay was performed according to the manufacturer’s instructions for use. 50 μL of Simplexas COVID-19 Direct Kit reaction mix (MOL4150) was added to the “R” well of the 8-well Direct Amplification Disc (DAD) followed by adding 50 μL of non-extracted nasopharyngeal swab (NPS) sample (collected in approximately 3 mL of Universal Transport Media (UTM, Copan)) to the “SAMPLE” well. Data collection and analysis was performed with LIAISON^®^ MDX Studio software. The assay targets two different regions of the SARS-CoV-2 genome, S gene and ORF1ab, differentiated with FAM and JOE fluorescent probes. An RNA internal control (Q670 probe) is used to detect RT-PCR failure and/or inhibition. The results interpretation algorithm for reporting a positive specimen requires only one of the two targets to be detected (S or ORF1ab gene).

### GenMark ePlex SARS-CoV-2 EUA panel

Testing with the ePlex SARS-CoV-2 panel was performed according to the manufacturer’s instructions for use. Briefly, after vortexing for 3-5 seconds, 200 μl of the primary NPS sample was aspirated into the sample delivery device (SDD) provided with the ePlex SARS-CoV-2 panel kit and vortexed once again for 10 seconds. The entire volume of the SDD was dispensed into the sample loading port of the SARS-CoV-2 test cartridge, followed by firmly pushing down on the cap to securely seal the sample delivery port. Each cartridge was bar-coded and scanned with the ePlex instrument and inserted into an available bay. Upon the completion of the assay run, the ePlex SARS-CoV-2 panel report was generated. The GenMark ePlex SARS-CoV-2 panel amplifies and detects the 2019-nCoV virus nucleocapsid (N) gene.

### Hologic Panther Fusion^®^ SARS-CoV-2 EUA

The Fusion SARS-CoV-2 assay was performed according to the manufacturer’s instructions for use. 500 µL of NPS specimens were lysed by transferring them to a Specimen Lysis Tube containing 710 µL lysis buffer. Input volume per sample for extraction is 360 µL. The Internal Control-S (IC-S) was added to each test specimen and controls via the working Panther Fusion Capture Reagent-S (wFCR-S). Hybridized nucleic acid was then separated from the specimen in a magnetic field. After wash steps, the elution step occurs and outputs 50µL of purified RNA. Then 5µL of eluted nucleic acid is transferred to a Panther Fusion reaction tube already containing oil and reconstituted mastermix. The Panther Fusion^®^ SARS-CoV-2 assay amplifies and detects two conserved regions of the ORF1ab gene in the same fluorescence channel. The two regions are not differentiated and amplification of either or both regions leads to a fluorescent ROX signal. The results interpretation algorithm for reporting a positive specimen requires only one of the two targets to be detected (ORF1a or ORF1b gene).

### Analytical Sensitivity

Limit of detection (LoD) was performed using a SARS-CoV-2 synthetic RNA quantified control (SARS-CoV-2 Standard) containing five gene targets (E, N, ORF1ab, RdRP and S Genes of SARS-CoV-2) from Exact Diagnostics (SKU COV019, Fort Worth, TX). A starting concentration of 200,000 copies/mL control was used to generate a dilution panel. The control was diluted in THE Ambion^®^ RNA Storage Solution (Catalog No. AM7001, ThermoFisher Scientific) and aliquoted for testing in order to obtain a maximum of 12 replicates at the following concentrations: 20,000, 2,000, 1,000, 500, 100, 50, and 5 copies/mL. The LoD was determined by two methods: Positive rate and Probit analyses. Positive rate was determined as the lowest dilution at which all replicates resulted positive with a 100% detection rate. The LoD by Probit was determined as the lowest detectable dilution at which the synthetic RNA quantified control (copies/mL) resulted positive with a 95% probability of detection. The final LoD was based on Probit analyses results and on each manufacturer’s claimed results interpretation algorithm.

### Discordant Analysis

Results were considered discordant when one molecular assay did not agree qualitatively (Detected or Not Detected) with the other three assay results. In such cases, molecular testing was repeated for the discordant assay.

### Statistical methods

The reference standard was established as a “consensus result” which was defined as the result obtained by at least three of the four molecular diagnostic assays. Percent sensitivity, specificity, positivity rate, Kappa, Probit, and two-sided (upper/lower) 95% confidence interval (CI) were calculated using Microsoft^®^ Office Excel 365 MSO software (Microsoft, Redmond, WA). The sensitivity was calculated as TP/(TP + FN) ⨯ 100, the specificity was calculated as TN/(TN + FP) ⨯ 100, where TP is true-positive results, FN is false-negative results, TN is true-negative results, and FP is false-positive results. Cohen’s kappa values (κ) were also calculated as a measure of overall agreement, with values categorized as almost-perfect (>0.90), strong (0.80 to 0.90), moderate (0.60 to 0.79), weak (0.40 to 0.59), minimal (0.21 to 0.39), or none (0 to 0.20) (12-13). Probit analyses were used for the copies/mL determination of the analytical sensitivity study. The dose-response 95th percentile (with 95% confidence interval [CI]) model was assessed using the Finney and Stevens calculations (14).

## Results

### Analytical Sensitivity

A serial dilution panel of SARS-CoV-2 control was tested to determine the LoD, defined as the minimum concentration with detection of 100% by positive rate and 95% by Probit analysis. The LoD established by percent positive rate ranged from 1,000 copies/mL by both the GenMark and the modified CDC assays to 50 copies/mL by the DiaSorin Molecular assay (**Table 2**). The LoD results were further subjected to Probit analysis. The 95% detection limit of the CDC assay was 779 ± 27 copies/mL for the N1 gene and 356 ± 20 copies/mL for the N2 gene. For the DiaSorin Molecular assay, the 95% detection limit was 39 ± 23 copies/mL for the S gene and 602 ± 28 copies/mL for ORF1ab. For the Hologic assay, the 95% detection limit was 83 ± 36 copies/mL for ORF1ab. Probit analysis could not be performed for the GenMark assay (**Table 2**). The final LoD, according to the assay results interpretation algorithm from each manufacturer, ranged from 1,000 copies/mL by the GenMark assay to 39 ± 23 copies/mL by the DiaSorin Molecular assay (**Table 2**).

**Table 2.** Summary of Limit of Detection results.

| Positive Rate % <sup>a</sup> |  |  |  |  |  |  |  | Probit<br>(95% CI)<sup>bc</sup> | Final LoD<sup>cd</sup> |
| --- | --- | --- | --- | --- | --- | --- | --- | --- | --- |
| No. of replicates detected at each dilution (copies/mL) |  |  |  |  |  |  |  | copies/mL | copies/mL |
| Molecular Assay |  | 2,000 | 1,000 | 500 | 100 | 50 | 5 |  |  |
| Modified CDC | N1 | 4/4<br>(100%) | **8/8**<br>**(100%)** | 7/10<br>(70%) | 1/10<br>(10%) | 0/8<br>(0%) | 1/4<br>(25%) | 779 ± 27 | 779 ± 27 |
|  | N2 | 4/4<br>(100%) | 8/8<br>(100%) | **10/10**<br>**(100%)** | 6/10<br>(60%) | 3/8<br>(38%) | 0/4<br>(0%) | 356 ± 20 |  |
| DiaSorin Molecular | S | 1/1<br>(100%) | 10/10<br>(100%) | 10/10<br>(100%) | 10/10<br>(100%) | **8/8**<br>**(100%)** | 0/4<br>(0%) | 39 ± 23 | 39 ± 23 |
|  | ORF1ab | 1/1<br>(100%) | **10/10**<br>**(100%)** | 8/10<br>(80%) | 4/10<br>(40%) | 1/8<br>(13%) | 0/4<br>(0%) | 602 ± 28 |  |
| GenMark | N | 10/10<br>(100%) | **9/9**<br>**(100%)** | 7/10<br>(70%) | 1/10<br>(10%) | 1/4<br>(25%) | 0/4<br>(0%) | N/A<sup>e</sup> | 1,000 |
| Hologic | ORF1ab | 3/3<br>(100%) | 9/9<br>(100%) | 12/12<br>(100%) | **12/12**<br>**(100%)** | 5/9<br>(56%) | 0/6<br>(0%) | 83 ± 36 | 83 ± 36 |

<sup>a</sup> The limit of detection by positive rate for each assay is highlighted in bold
<sup>b</sup> CI, confidence interval
<sup>c</sup> ±, upper/lower 95%
<sup>d</sup> The final LoD was based on each manufacturer's results interpretation algorithm
<sup>e</sup> Not applicable

### Clinical performance of four EUA SARS-CoV-2 (COVID-19) molecular assays

Following testing of 104 retrospective clinical specimens, the modified CDC, DiaSorin Molecular, and Hologic EUA molecular assays demonstrated a sensitivity of 100% (51/51), while the GenMark ePlex SARS-CoV-2 EUA panel showed a sensitivity of 96% (49/51). A specificity of 100% (53/53) was observed for GenMark and DiaSorin Molecular, while percent specificities ranged from 98% (52/53) for CDC to 96% (51/53) for Hologic (**Table 3**).

**Table 3.**
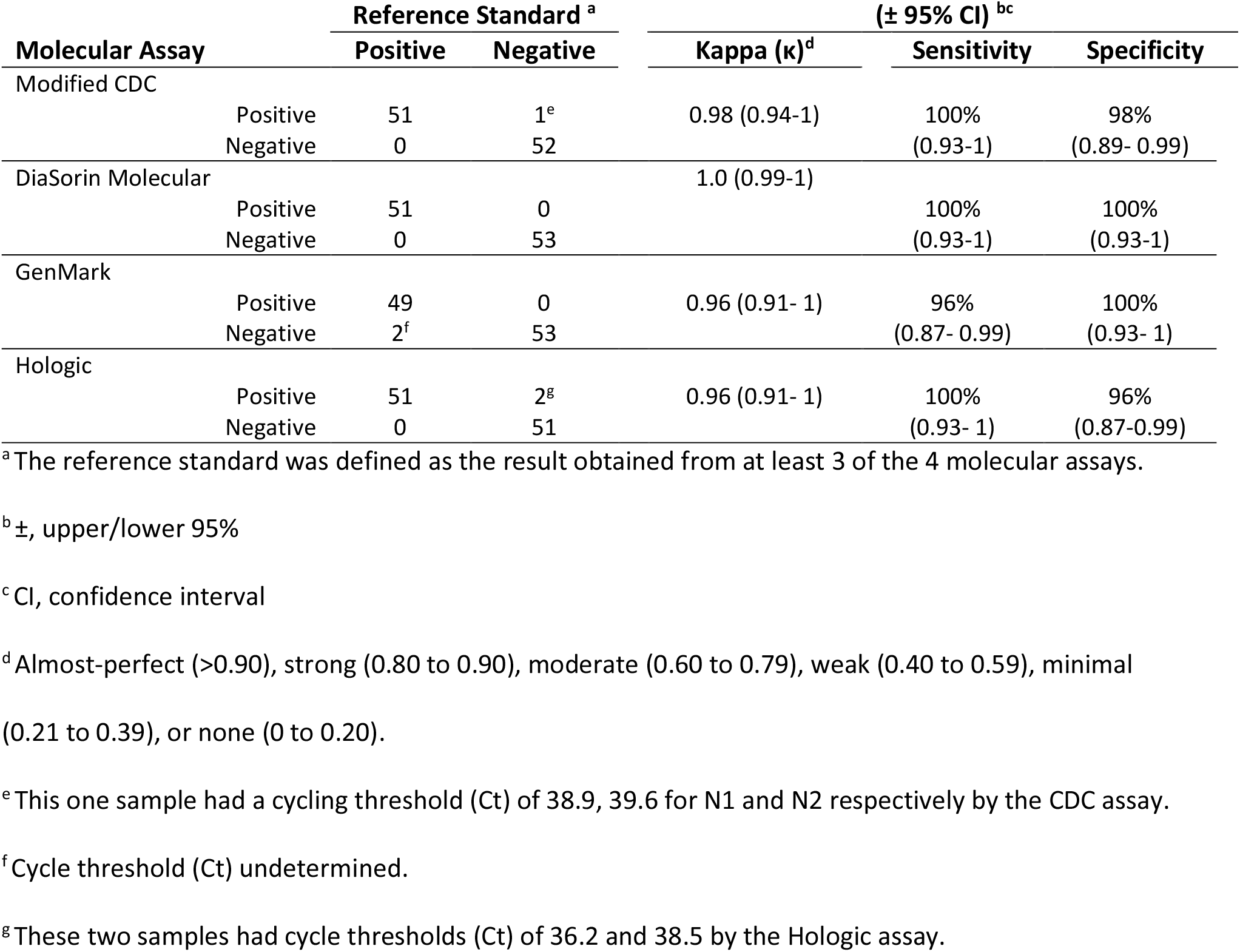
Clinical performance comparison of four EUA molecular assays for the detection of SARS-CoV-2 in nasopharyngeal swab specimens (n = 104).

Details for discordant sample analysis are shown in **Table 4**. A total of five discordant samples were found among three out of the four platforms. One false positive sample (NW-104) had Ct values of 38.9 and 39.6 for N1 and N2 genes, respectively, on initial testing by the modified CDC assay. After repeating extraction and retesting, the sample was determined to be negative. Two samples (NW-97 & NW-99) were considered false negative by GenMark but positive by the other three methods. After reprocessing and retesting, the GenMark assay was able to detect both samples as positive. Two additional false positive samples (NW-83 & NW-85) were found by the Hologic assay; original samples were retested and were found to be positive and negative, respectively. Following retesting of the five discordant samples, the GenMark ePlex SARS-CoV-2 EUA panel showed an improvement of sensitivity to 100% (51/51). Additionally, a 100% specificity (53/53) was obtained for the CDC assay, while Hologic improved to 98% (52/53) (**Table 4**).

**Table 4.** Details of discordant sample analysis.

| Sample ID | Reference Standard | SARS-CoV-2 Molecular Assay Results (Ct)<sup>ab</sup> |  |  |  | Comment |
| --- | --- | --- | --- | --- | --- | --- |
|  |  | Modified CDC (N1/N2) | DiaSorin Molecular (S/ORF1ab) | GenMark | Hologic |  |
| NW-83 | NEG | NEG | NEG | NEG | **POS** (36.2) | Sample was repeated by Hologic and determined POS |
| NW-85 | NEG | NEG | NEG | NEG | **POS** (38.5) | Sample was repeated by Hologic and determined NEG |
| NW-97 | POS | POS (35.5/ 34.5) | POS (31.9/31.8) | **NEG** | POS (35.0) | Sample was repeated by GenMark and determined POS |
| NW-99 | POS | POS (35.3/35.0) | POS (29.3/29.9) | **NEG** | POS (33.0) | Sample was repeated by GenMark and determined POS |
| NW-104 | NEG | **POS** (38.9/39.6) | NEG | NEG | NEG | Sample was repeated by CDC and determined NEG |

<sup>a</sup> Discordant sample results are highlighted in bold
<sup>b</sup> Ct, Cycle threshold

### Workflow evaluation

Hands-on time (HOT), run time, and overall turnaround time (TAT) to results were assessed for all pre-analytical, analytical, and post-analytical steps for all four platforms. Results of the workflow assessment are shown in **Table 5**. The HOT ranged between all platforms. The longest hands-on time was the Hologic assay at ∼2 hours followed by the modified CDC assay with ∼1 hour and 30 minutes. Very comparable HOT was found for DiaSorin Molecular and GenMark with a range of 16 minutes and 12 minutes, respectively. The run time averaged 90 minutes for modified CDC, DiaSorin Molecular, and GenMark. Hologic was the exception with 4 hours with 35 mins of run time (**Table 5**). Overall turn-around time assessment, from sample to results, showed DiaSorin Molecular with the least overall turn-around time to results, followed by GenMark, modified CDC assay and Hologic with the greatest overall time (**Table 5**).

**Table 5.** Throughput and workflow evaluation for four EUA molecular SARS-CoV-2 assays.

|  | SARS-CoV-2 Molecular Assay |  |  |  |
| --- | --- | --- | --- | --- |
|  | 2019-Novel Coronavirus<br>(2019-nCoV) | Simplexa™ COVID-19<br>Direct | ePlex SARS-CoV-2 Test | SARS-CoV-2 Assay |
| **Manufacturer** | Modified CDC | DiaSorin Molecular | GenMark | Hologic |
| **Throughput**<br>**(samples per run)** | 24 | 8 per disc | 6 per tower | 120 |
| **Input volume per**<br>**sample/Elution**<br>**volume** | 110 µL/110 µL | 50 µL/NA | 200 µL/NA | 360 µL/50µL |
| **Hands-on Time**<br>**(per run)** | ~1.5 hr | < 16 min | < 12 min | ~2.0 hrs |
| **Assay Run Time** | ~90 min | ~ 90 min | ~ 90 min | ~4 h 35 min |
| **User Results**<br>**Interpretation** | Yes | No | No | No |
| **Overall Turn-**<br>**around Time** | ~3 hr | ~1.8 hr | ~1.7 hr | ~ 6.6 hr |

## Discussion

In this study, we compared four different platforms for the detection of SARS-CoV-2 in patient specimens collected during March and April of the 2020 COVID-19 outbreak in the United States. We were able to make several observations, including LoD, overall workflow comparisons, and how each test performed in a head-to-head clinical comparison. Accurate and actionable results have been at the core of medical decision-making during this current outbreak, both in the inpatient and outpatient setting. For hospitalized patients, results are critical for clinical management as well as infection control and cohorting for bed management. Likewise, results are just as critical in the outpatient setting as the basis for social distancing measures to slow the spread of infection. To that end, false negative results are particularly troubling, since they inevitably lead to more exposures. Turnaround time is also critical for allocation of limited resources, such as the limited availability of isolation rooms and real-time cohorting decisions. In addition, healthcare workers need rapid results to ensure they are not exposing patients whom they are treating. Moreover, levels of personal protective equipment (PPE) required by health care professionals also vary depending on whether a patient is COVID-19 positive, requiring a rapid TAT to preserve precious resources. Considering the transmissibility of SARS-CoV-2, which has recently been estimated to have a basic reproduction number (R0) of 2.2, meaning that on average, each infected person can spread the infection to an additional two persons, a false negative result can be devastating (15, 16). This is especially true in vulnerable patient populations such as the elderly (especially people living in a nursing home or long-term care facility), immunocompromised, and in people with pre-existing medical conditions (17, 18).

Our data suggest that all four PCR methods yielded comparable results (κ ≥ 0.96); however, we did observe a notable difference in the sensitivity of the methods during this large-scale evaluation of EUA *in vitro* diagnostic assays. Our study showed that the DiaSorin Molecular and Hologic Fusion assays out-performed both the modified CDC and GenMark assays when it came to overall LoD, with GenMark having the overall highest LoD of all four platforms evaluated. DiaSorin Molecular had the lowest LoD (39 ± 23 copies/mL), closely followed by Hologic (83 ± 36 copies/mL). The modified CDC assay showed a final LoD, of 779 ± 27 copies/mL based on the results interpretation algorithm. It is worth mentioning that this assay requires both targets to be fully detected, thus clinical samples falling in this concentration range would be identified and repeated, potentially generating additional turnaround time and laboratory labor. In contrast, GenMark could only detect 100% of replicates at 1,000 copies/mL, and was not able to reliably detect replicates below 1,000 copies/mL, thus patient specimens below this concentration range could potentially be missed. One important limitation to mention is that sensitivity using Probit analysis could not be calculated for GenMark since Ct values are not available as part of the ePlex system result interpretation.

The clinical correlation was also consistent with LoD findings, where both the DiaSorin Molecular and Hologic assays had 100% sensitivity and detected all specimens deemed positive by the consensus standard (interpretation of three of four evaluated assays as “gold standard”), whereas GenMark missed two positive specimens (which were subsequently detected by GenMark upon repeat). DiaSorin Molecular and GenMark showed 100% specificity, while Hologic and the CDC assay initially had two and one discordant results, respectively. Repeat testing of these three specimens showed that for Hologic, NW-83 repeated as positive a second time and was therefore potentially a false positive and NW-85 was negative upon repeat, meaning this result could have previously been a false positive as well. The continued discordant result from NW-83 could potentially be attributed to specificity issues, since the assay did not prove to be the most sensitive assay among those used in the study, with DiaSorin Molecular showing a slightly lower LoD. Repeat testing of NW-104 on the modified CDC assay was negative. Considering the LoD of the modified CDC assay, coupled with the fact that both DiaSorin Molecular and Hologic resulted NW-104 as negative, this was likely a false-positive result. While all 4 assays could reliably detect most patients in our study, GenMark lacked sensitivity, initially missing two low-level positive specimens, and this could easily have impacted patient diagnosis by missing true positive patients.

When it comes to the hands-on and turnaround time of the four assays in this study, the throughput and workflow evaluation are clearly shown in **Table 5** and are based on lab technologist experience in our laboratory. As a routine real-time RT-PCR assay, the modified CDC requires nucleic acid extraction, master mix preparation and PCR setup, standard PCR amplification, as well as interpretation of the results. This involves several manual steps, needing about 1 hr 30 min hands-on time and an approximate overall turnaround time of 3 hours. The DiaSorin Molecular and GenMark assays have comparatively similar hands-on time and turnaround time, based on processing 8 samples per disc on the DiaSorin LIAISON MDX and 6 cartridges per tower in the GenMark ePlex. Clinical laboratories may decide to purchase additional instruments to allow for testing of more samples at a time in order to satisfy patient testing volume requirements. The Hologic Panther Fusion platform has more of an automated workflow, with five samples processed at a time after loading. The sample to answer time for the first five samples is 2 hours and 40 min, followed by 5 results every 5 minutes after loading 120 samples; the total assay run time for 120 specimens is approximately 4 hours and 35 min. It is also important to note that the Hologic platform has longer hands-on time, since the technologist has to load the primers, probes, and other consumables and the fact that 120 clinical samples have to be manually transferred to Sample Lysis Buffer tubes. These steps, especially the pipetting of the specimen into the lysis tube, can be somewhat labor intensive and time consuming, bumping the overall turnaround time for 120 specimens closer to the 7 hour mark. It is important to emphasize that each platform has their advantages. For workflow, TAT, and ease of use, the three sample-to-answer platforms (DiaSorin Molecular, Hologic, GenMark) out-performed the modified CDC assay, which is a manual assay requiring many steps, specialized personnel, and separate areas for processing and performing the test. The Hologic platform is more appropriate for high-volume testing, while the DiaSorin Molecular and GenMark systems both work well in an environment where rapid results and lower to moderate testing volumes are required.

This study has several limitations that should be mentioned. First, this was a single center study and the majority of the specimens were frozen after initial testing on the GenMark assay. While these limitations are present, they have been minimized by the fact that GenMark assay (which was the least sensitive platform in the analysis) actually had a potential competitive advantage, since it was the assay initially performed on fresh specimens. Second, while the number of specimens included in the clinical correlation was only 104, the patient samples spanned the entire range of clinical positives and reflected our overall true positivity rate, which was between 50-60% during this time period of the COVID-19 outbreak.

In summary, we have evaluated four molecular *in vitro* diagnostic assays for the qualitative detection of SARS-CoV-2 in nasopharyngeal specimens. The data from our evaluation suggest that the modified CDC, DiaSorin Molecular, Hologic and GenMark assays performed similarly (κ ≥ 0.96) and that all but the CDC assay can function in a sample-to-answer capacity. The GenMark assay, however, was less sensitive and had a higher LoD than both the DiaSorin Molecular and Hologic assays. When considering the design of all four assays, differences that could affect assay performance could include characteristics such as input volume of initial specimen, RNA purification and elution volume differences, and overall differences in gene targets. The DiaSorin Molecular platform has lower testing volume capability compared to the Hologic assay (8 specimens/disc run vs. 120 specimens loaded at once), but has a faster TAT and less reagent/sample preparation. All of these parameters, along with patient care needs, may assist clinical laboratories to identify and choose the correct testing platform that best fits their needs for the diagnosis of patients infected with this novel human coronavirus.

## Data Availability

numbers from raw data are summarized in the manuscript to determine LoD, sensitivity, specificity, etc.

## Acknowledgements and Disclosures

We would also like to thank Diasorin Molecular LLC. for providing the reagents used in this study. Gregory Berry has previously given education seminars for Hologic, Inc., and received an Honorarium.

